# A novel patient with inherited TNF deficiency and tuberculosis

**DOI:** 10.64898/2026.11.09.26366865

**Authors:** Narelle Keating, Mehrnaz Mesdaghi, Zahra Chavoshzadeh, Abdollah Karimi, Mohammad Saberi, Sinem Ünal, Vincent Meynier, Camille Soudée, Candice Lainé, Abbas Solgi, Samin Sharafian, Anne-Sophie L’Honneur, Mélanie Migaud, Yoann Seeleuthner, Aurélie Cobat, Laurent Abel, Vivien Béziat, Jérémie Rosain, Jacinta Bustamante, Jean-Laurent Casanova, Stéphanie Boisson-Dupuis

## Abstract

Two otherwise healthy patients from Colombia, now aged 30 and 38 years, developed pulmonary tuberculosis due to inherited, complete TNF deficiency at the ages of 19 and 18 years. We report here a previously healthy Iranian patient with inherited complete TNF deficiency who was diagnosed with both pulmonary and extrapulmonary TB between 10 and 15 years of age. The homozygous *TNF* variant (c.673C>T) introduces a premature stop codon (TNF p.Q225*) resulting in losses of expression and function. Despite normal myeloid and lymphoid subset development, the patient’s primary leukocytes do not secrete detectable amounts of TNF. As in the TNF-deficient Colombian patients, the respiratory burst of GM-CSF-derived macrophages is impaired. Our findings expand the spectrum of TB phenotypes in patients with TNF deficiency and confirm that TNF is essential for immunity to *M. tuberculosis* in humans but apparently redundant against many other pathogens, including weakly virulent mycobacteria.

## Introduction

Tuberculosis (TB) is most frequently triggered by *Mycobacterium tuberculosis (M. tb)*^1^. A quarter of the world’s population is latently infected with *M. tb*^1^. In 5-10% of cases, infection progresses to active TB, which remains the leading infectious cause of death worldwide^1^. Discoveries of the genetic etiologies underlying Mendelian susceptibility to mycobacterial disease (MSMD), characterized by susceptibility to weakly virulent mycobacteria and the Bacillus Calmette-Guérin (BCG) vaccine (an attenuated live strain of *Mycobacterium bovis*)^2^, were instrumental in demonstrating that TB can also be a monogenic disorder in some patients^3,4^. Indeed, genetic defects disrupting interferon-gamma (IFN-γ) immunity have been reported in patients with TB. Biallelic genotypes involving rare variants of *TNF*^5^, *LY9*^6^*, ITK*^7^*, IL12RB1*^8^*, IL23R*^9^*, SPPL2A*^10^ and *PDCD1*^11^ and the common variants *IL23R R381Q*^9^ and *TYK2 P1104A*^12^ have all been reported to underlie TB. Human genetics studies have clearly shown that the protection against TB provided by host immunity is dependent on an intact IFN-γ circuit, including the molecules inducing IFN-γ and those induced by IFN-γ, to eliminate the mycobacteria within macrophages. Tumor necrosis factor (TNF) is a pro-inflammatory cytokine produced by macrophages in response to IFN-γ; it drives inflammation, promotes cell death and activates the respiratory burst mediated by the production of reactive oxygen species (ROS) during infection^5,13^. IFN-γ can therefore drive ROS production both directly and indirectly, by upregulating the transcription of the NADPH oxidase components involved in the respiratory burst^14^ and inducing the production of TNF^15^.

In this context, we previously described two patients with TB from the same Colombian family with inherited, complete TNF deficiency^5^. TNF has been implicated in host defense against multiple pathogens in mice^16^, but these two patients presented with recurrent pulmonary TB, despite broad pathogen exposure, including BCG vaccination. They were homozygous for a private *TNF* variant underlying complete TNF loss-of-function (LOF)^5^. They developed TB at the ages of 19 and 18 years. They are now 30 and 38 years of age, respectively, in complete remission, and healthy. TNF deficiency weakened antimycobacterial defense at the cellular level, by impairing the respiratory burst in GM-CSF-matured and alveolar macrophages^5^, but not in neutrophils. This observation is consistent with rare *CYBB* variants selectively impairing ROS production in macrophages and leading to MSMD, a phenotype largely restricted to mycobacterial disease^17–19^. By contrast, patients with chronic granulomatous disease (CGD) have a broad clinical phenotype due to the impairment of ROS production in all phagocytes^20^. We searched for new patients with autosomal recessive (AR) TNF deficiency, to expand our allelic, clinical and cellular understanding of TNF deficiency.

## Results

### Case presentation

We studied a female patient (P) born to consanguineous parents from Iran **(Figure 1A)**. She was diagnosed with miliary pulmonary TB (between 10-15 years of age) with dissemination to the left pelvic bone, presenting with fever, fatigue, muscle pain, and an unexplained loss of about 3 kg of body weight. Acid-fast microscopy on a sputum sample revealed the presence of *M. tb*., and a tuberculin purified protein derivative (PPD) skin test with 5 tuberculin units resulted in an induration of 25 mm. These tests were also performed on the patient’s parents and sibling, yielding negative results. CT scans of the patient’s lungs were indicative of miliary TB **(Figure 1B)**. At the time of diagnosis, immunological profiling showed the patient to have essentially normal proportions of leukocytes. The patient had a leukocyte count of 8800/mm^3^, with the proportions of neutrophils (36%), eosinophils (5.3%), and lymphocytes (45%) all within the normal range. The proportion of monocytes was slightly high, at 12%. Acute-phase reactant levels were high, with a C-reactive protein (CRP) concentration of 63 mg/liter. The patient received a three-month course of anti-TB treatment with a five-drug regimen consisting of isoniazid, rifampin, ethambutol, levofloxacin, and clarithromycin, followed by nine additional months of treatment with isoniazid and rifampin, leading to complete recovery. The patient was vaccinated with BCG at birth, without adverse effects, and none of her relatives had any history of TB. The patient was also diagnosed with hypothyroidism and is currently being treated with levothyroxine. She was otherwise healthy and is HIV-negative. No autoantibodies against IFN-γ, IL-12, IL-23 or TNF were detected in the patient’s serum. No other unusual infections were reported. Serological testing **(Table 1)** and VirScan analysis **(Figure 1C)** of the patient confirmed normal antibody repertoires against common pathogens, such as human coronavirus OC43, cytomegalovirus, and norovirus, demonstrating intact T cell and B cell functions for seroconversion after vaccination and immunity to common infections.

**Figure 1:**
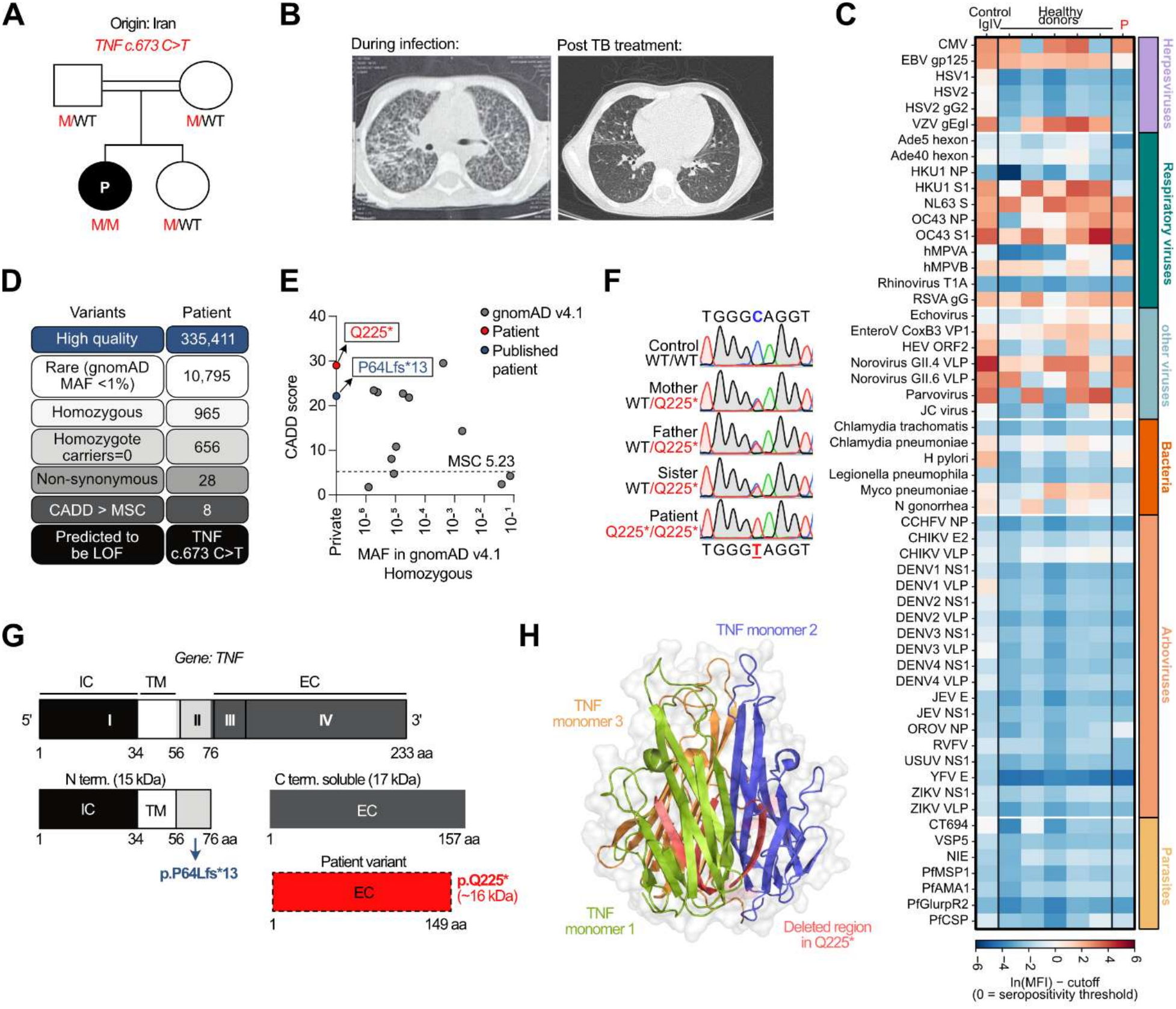
TNF deficiency detected in a patient with miliary pulmonary tuberculosis and osteomyelitis. **A:** Pedigree and familial segregation of the *TNF* variant (c.673 C>T; p.Q225*). **B:** CT scan of the patient’s lungs pre– and post-TB treatment. **C:** VirScan analysis of the patient and five healthy controls. Pooled human plasma used for IVIg (Privigen® CSL Behring AG) was used as an additional control. Zero is the seropositivity threshold; red is above, blue below. **D:** Whole-exome sequencing triage. CADD=combined annotation-dependent depletion, MSC=mutational significance cutoff. **E:** CADD vs. minor allelic frequency (MAF) graph for homozygous *TNF* variants from gnomAD v4.1, highlighting the position of the patient’s allele and of the previously published p.P64Lfs*13 variant. **F:** The alleles present in the mother, father and sister of the patient and in the patient herself were confirmed by Sanger sequencing. **G:** Schematic representation of *TNF*, with the exons indicated in roman numerals. Protein domains are annotated above the gene. Aa=amino acids, IC=intracellular, TM=transmembrane, EC=extracellular. **H:** Cartoon representation of trimeric TNF (aa77-233). Single monomers are depicted in purple, orange or green, with the deleted regions shown in red. PDB: 1TNF.

**Table 1:** Viral serology for the TNF-deficient patient. CMV: cytomegalovirus; EBNA: Epstein-Barr virus nuclear antigen 1; HAV: hepatitis A virus; HSV: herpes simplex virus; VCA: viral capsid antigen; VZV: varicella zoster virus.

| **Parameters** | **Patient** | **Threshold** |
|---|---|---|
| IgG anti-HSV-1 | 0.09 | >1.1 |
| IgG anti-HSV-2 | <0.5 | >1.1 |
| IgG anti-VZV | 26.96 | >165 |
| IgG anti-EBNA | <3 | >20 |
| IgG anti-VCA | <10 | >40 |
| IgG anti-CMV | 96.6 | >14 |
| IgG anti-HAV | 0.93 | <0.9 |
| IgG anti-parvovirus B19 | <0.1 | >1.1 |
| IgG anti-measles | <5 | >16 |
| IgG anti-mumps | <5 | >11 |
| IgG anti-rubella | 5.33 | >10 |

### Homozygosity for a private variant of *TNF*

Whole-exome sequencing (WES) revealed homozygosity for a private variant in exon 4 (c.673C>T) of *TNF* resulting in a premature stop codon (TNF p.Q225*) **(Figure 1D)**. The patient had a homozygosity rate of 9.7%, confirming that her parents were, indeed, consanguineous. *In silico* predictions for TNF p.Q225* suggested that it would be highly deleterious with a combined annotation-dependent depletion (CADD) score of 29, above the mutation significance cutoff of 5.23 for *TNF* and the CADD score for the previously published variant (TNF p.P64Lfs*13)^5^ **(Figure 1E)**. Furthermore, TNF p.Q225* had never been reported in any public databases (gnomADv4.1) and was absent from our in-house database of >25,000 exomes **(Figure 1E)**. Sanger sequencing confirmed that the patient was homozygous for c.673C>T and that the patient’s mother, father, and sister were heterozygous for the same variant **(Figure 1F)**. TNF is produced as a transmembrane protein, the extracellular domain of which is cleaved to form a soluble trimer that binds and activates TNF receptors^21^. The premature stop codon occurred within the soluble fragment of the protein **(Figure 1G)** and eliminated the final beta sheet in its entirety **(Figure 1H).** We therefore hypothesized that TNF p.Q225* would impair TNF function, accounting for the patient’s susceptibility to TB.

### TNF p.Q225* confers a complete functional deficiency in an overexpression system

We assessed the impact of the variant on the production of the corresponding mRNA and protein *in vitro* by generating cDNA constructs for the expression of TNF p.Q225* in HEK293T cells. As controls, we included the previously described TNF p.Y163H and TNF p.A221R variants^22^ (encoding proteins that do not bind TNFR1 and TNFR2), the TNF p.V77* variant (which cannot be cleaved to generate a soluble fragment^5^), and the previously reported TNF p.P64Lfs*13 variant^5^. In a transient overexpression system in HEK293T cells, the mRNA encoding TNF p.Q225* was present in similar amounts to the mRNAs encoding WT TNF and p.P64Lfs*13 **(Figure 2A)**. However, on immunoblot analysis, the TNF p.Q225* protein was undetectable in both the cell lysate and supernatants harvested from cells transfected with TNF p.Q225* **(Figure 2B)**. As expected, p.V77* and p.P64Lfs*13 were also undetectable, and p.Y163H and p.A221R were detected but present in smaller amounts than WT TNF. Importantly, the epitope of the anti-TNF antibody used in these analyses occurs upstream from the p.Q225* variant. These results suggest that the patient’s variant results in a loss of TNF. We then investigated the functional impact of TNF p.Q225* in an NF-κB luciferase assay on HEK293T cells. Soluble TNF was generated by transiently transfecting HEK293T cells with the WT and mutant TNF alleles, or with empty vector (EV) used as a negative control. The supernatant was harvested 24 h later. HEK293T cells transiently transfected with NF-κB–Firefly and Renilla luciferases (as a transfection control) were then stimulated with these supernatants, or with commercially available recombinant human TNF, overnight. TNF–induced NF-κB activity was detected in cells simulated with WT TNF but was absent from cells stimulated with TNF p.Q225* **(Figure 2C),** confirming that TNF p.Q225* was, indeed, a loss-of-function (LOF) allele.

**Figure 2:**
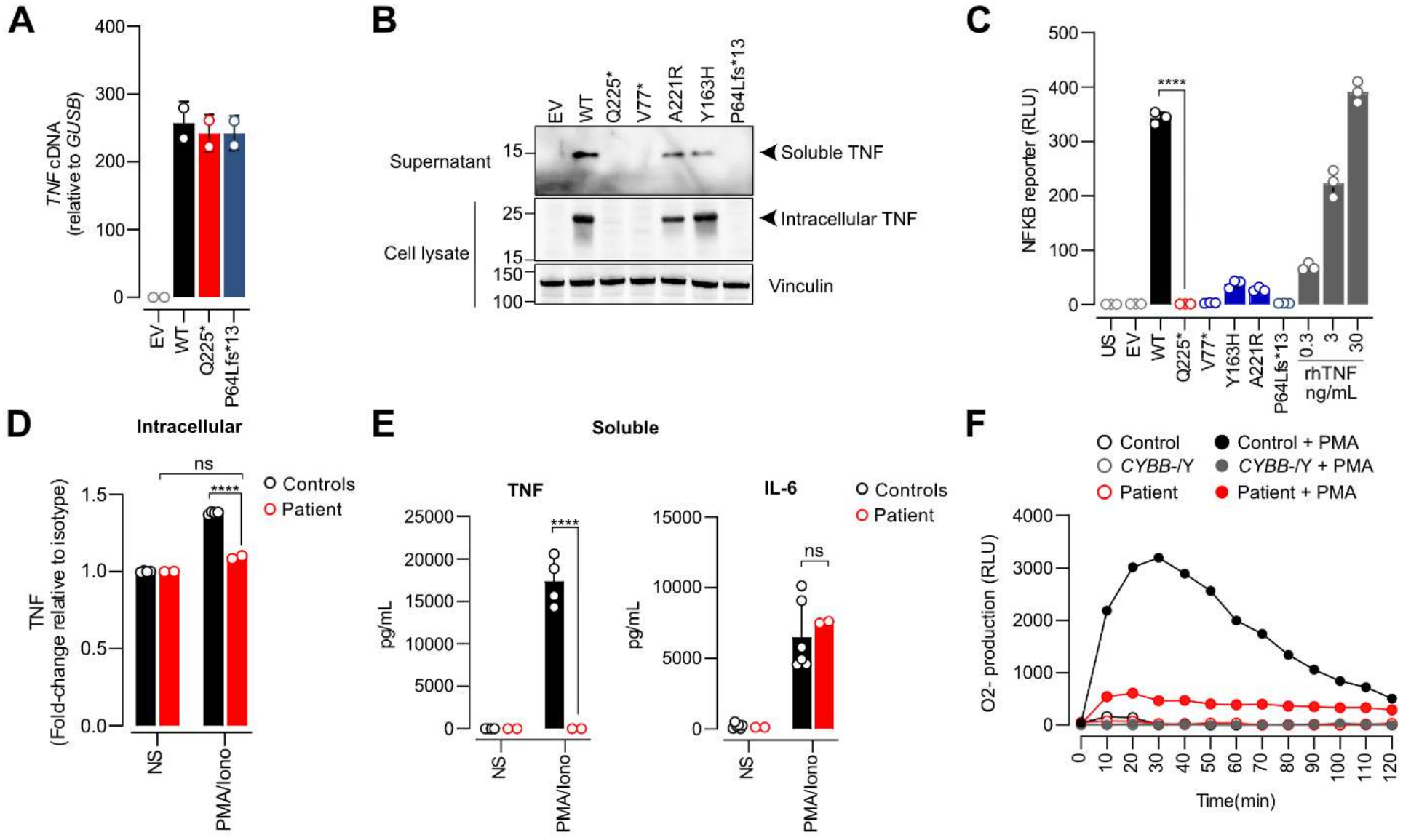
TNF p.Q225* is a loss-of-expression and loss-of-function allele. **A**: HEK293T cells were transiently transfected with EV, WT or mutant TNF constructs and analyzed by RT-PCR. Samples were run in technical duplicates and data are presented as the mean ± SD. **B:** HEK293T cells were transfected as in **A**. Supernatants were harvested and the cells were lysed for SDS-PAGE and immunoblot analysis with anti-TNF and anti-Vinculin antibodies (as a loading control). Representative results from three independent experiments are shown. **C:** HEK293T cells were transiently transfected with NF-κB–Firefly and Renilla luciferase plasmids. Cells were stimulated, 24 h post-transfection, with recombinant human TNF (0.3-30 ng/mL) or with supernatants harvested from transfected-HEK293T cells. Data are presented as relative luciferase activity (RLU; Firefly to Renilla luciferase signal ratio) and expressed as a fold-change relative to unstimulated (US) cells. Samples were run in technical triplicates and data are presented as the mean ± SD. **D-E:** Peripheral blood mononuclear cells (PBMCs) were isolated from whole blood obtained from two healthy controls and the TNF p.Q225* patient and were stimulated for 6 h with PMA/ionomycin. Intracellular TNF expression was assessed by flow cytometry **(D)**, and soluble TNF and IL-6 expression was assessed in a Legend-plex assay **(E).** NS=not stimulated. **F:** Production of O_2_ ^−^, shown in relative luminescence units (RLU), by GM-CSF-matured MDMs from a healthy control, a gp91*^phox^*CYBB-deficient patient and the patient with TNF p.Q225*, after stimulation with PMA. This experiment was performed once, due to limited availability of patient material. The statistical significance of differences between groups was assessed in unpaired *t*-tests. ****=*p* < 0.0001.

### The patient’s leukocytes lack functional TNF

We assessed the production and function of TNF p.Q225* in primary patient leukocytes directly, by isolating peripheral blood mononuclear cells (PBMCs) from the whole blood of the patient and two healthy donors as controls. Flow cytometry staining for TNF on phorbol 12-myristate 13-acetate (PMA)/ionomycin-stimulated PBMCs detected intracellular TNF in the control cells but not in the cells of the patient **(Figure 2D)**. Determinations of soluble TNF in the supernatant of PMA/ionomycin-stimulated cells showed an absence of TNF secretion by the patient’s cells, whereas normal amounts of IL-6 were detected **(Figure 2E)**. We then assessed ROS production in GM-CSF-matured monocyte-derived macrophages (MDMs) in a luminol assay. In this assay, *CYBB*^-/Y^ MDMs were used as a control for defective ROS production. ROS production in response to stimulation with PMA was impaired in patient-derived GM-CSF MDMs relative to those derived from a healthy control **(Figure 2F)**. Importantly, a dihydrorhodamine (DHR) test performed on whole blood from the patient revealed no change in ROS production in neutrophils (data not shown). Thus, the cells of the patient did not produce or secrete functional TNF and ROS production was impaired in her GM-CSF-derived macrophages, accounting for her susceptibility to TB.

### Normal leukocyte development

We then investigated the effect of TNF deficiency on leukocyte development in this patient. Complete blood-cell counts were normal (data not shown). Five years post-TB recovery, we obtained whole blood from the patient and used mass cytometry (CyTOF®: Cytometry by Time-Of-Flight) to investigate the various PBMC subsets of the patient, comparing them with those of healthy controls matched with the patient for age and sex. The patient had slightly lower proportions and absolute counts of total and naïve CD4^+^ T cells, but higher proportions (but not counts) of effector memory and effector memory re-expressing CD45RA (TEMRA) CD4^+^ T cells **(Figure 3A)**. She had similar proportions of central memory CD4^+^ T cells to the controls **(Figure 3A)**. The frequencies of γδ T cells were unaffected by TNF deficiency **(Figure 3B)**, whereas the frequencies and counts of mucosal-associated invariant T (MAIT) cells were higher than in controls **(Figure 3C)**. The patient had similar proportions of CD8^+^ T cells, naive, central memory and effector memory subsets, and higher proportions, but not absolute counts, of TEMRA CD8^+^ T cells **(Figure 3D)**. The proportions of the T helper, natural killer (NK) cell, B-cell, monocyte and dendritic-cell (DC) subsets were all within the normal range **(Figure 3E-I)**. Overall, these data suggest that complete TNF deficiency did not alter the development of the major myeloid and lymphoid subsets in the blood.

**Figure 3:**
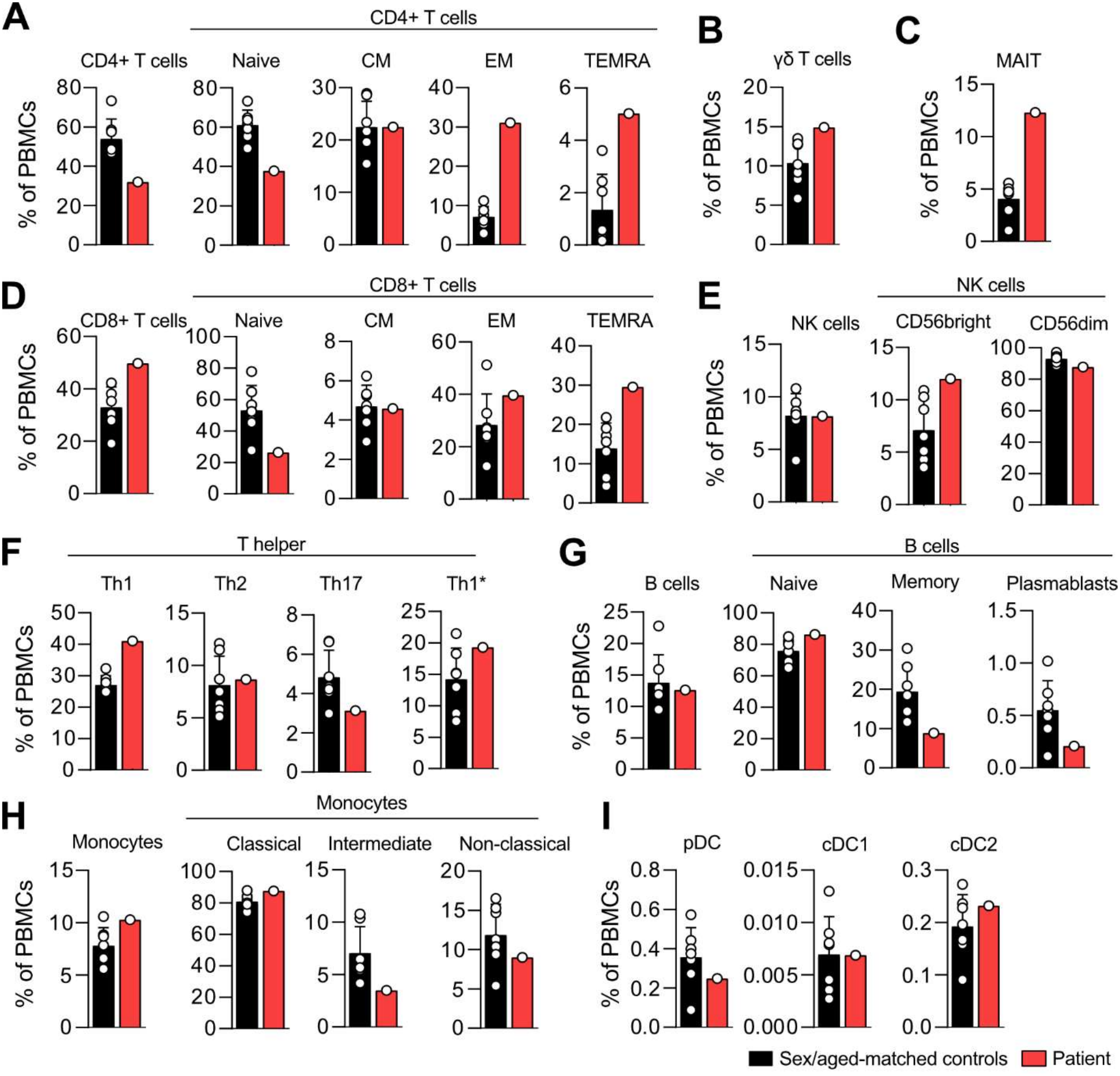
Deep immunophenotyping by CyTOF for the TNF-deficient patient. Frequency of **(A)** CD4^+^ T cells and their subsets, **(B)** TCR γδ^+^ T cells, **(C)** mucosal-associated invariant T cells (MAIT), **(D)** CD8^+^T cells and their subsets, **(E)** NK cells and their subsets, **(F)** T helper (Th) cells and their subsets, **(G)** B cells and plasma cells, **(H)** monocytes and their subsets, and **(I)** dendritic cell (DC) subsets, in healthy controls matched with the patient for age and sex (*n* = 10) and the patient, as determined by CyTOF. CM= central memory, EM=effector memory, TEMRA=effector memory re-expressing CD45RA, pDC=plasmacytoid dendritic cells, cDC=conventional dendritic cells.

## Discussion

We report a new patient with inherited TNF deficiency as a genetic etiology of TB, confirming the importance of TNF as a non-redundant component of human immunity to *M. tb*. The defects observed in this patient are consistent with our findings for other TNF-deficient patients^5^, in whom impaired TNF signaling leads to defective ROS production in GM-CSF–matured MDMs *ex vivo*, with broader myeloid and lymphoid differentiation remaining intact. This dissociation between the preservation of hematopoietic development and selective effector dysfunction highlights a specialized role for TNF in macrophage antimicrobial programming rather than in immune lineage specification. Collectively, these data support a mechanistic model in which TNF is crucial for the optimization of macrophage bactericidal activity via ROS-dependent pathways to provide essential protection against *M. tb* infection. Our findings for this patient extend the genetic and clinical field of TNF deficiency-associated TB to new geographic and ethnic backgrounds, adding an Iranian individual to the previously reported Colombian cases, thereby increasing the generalizability of TNF deficiency as a monogenic cause of TB. A highly consistent immunological phenotype was observed in all the reported patients: preserved resistance to the agent used for BCG vaccination and to a range of other common intracellular pathogens, including environmental and atypical mycobacteria, but with selective susceptibility to *M. tb*. This pattern suggests that TNF is not broadly required for host defense against most pathogens, instead playing a more specific role in optimizing antimycobacterial effector mechanisms, particularly those linked to ROS generation in GM-CSF–matured macrophages. Unlike the previously reported Colombian patients, who had primarily pulmonary disease, this patient had disseminated TB, with lesions in her left pelvic bone.

TB is the most lethal human infectious disease worldwide^1^, reflecting the ability of *M. tb* to exploit vulnerabilities in host immunity. It is, therefore, unsurprising that any impairment of ROS production by TNF deficiency, which is crucial for macrophage function, renders the host susceptible to severe TB. Conversely, it is surprising that TNF deficiency results in a selective susceptibility to TB disease and not to other pathogens, including weakly virulent mycobacteria. The data presented here and elsewhere show that the impairment of ROS production in TNF-deficient macrophages is not as severe as that observed in the cells of CGD patients (those with gp91***^phox^*** CYBB deficiency, for example)^17,20^. It is possible that the ‘milder’ impairment in ROS production allows the host to maintain immunity to bacteria other than *M. tb*. Five TNF antagonists have been developed and approved for clinical use (etanercept, infliximab, adalimumab, certolizumab and golimumab); they are mostly used to treat Crohn’s disease and rheumatoid arthritis^23^. A strong association has been found between anti-TNF therapy and an increase in susceptibility to opportunistic infections with intracellular pathogens, such as Whipple’s disease^24^ and listeriosis^25^, together with an increase in the risk of latent TB infection reactivation^26^. Additionally, TNF-blockade in humans poses serious risks of endemic mycoses^27–29^, including histoplasmosis, coccidioidomycosis, and blastomycosis, largely through impaired granuloma formation and macrophage activation. Patients with genetic defects depriving them of TNF may be vulnerable to these infections.

## Acknowledgments

We thank the patient, her relatives and the physicians for participating in this study; Yelena Nemirovskaya, Lazaro Lorenzo, Candace Clift, Maya Chrabieh, and Mark Woollett for administrative support; and all of the members of the Laboratory of Human Genetics of Infectious Disease for discussions.

## Author Contributions

Conceptualization: JR, JB, JLC, SBD.

Methodology: All authors.

Formal analysis and investigation: All authors.

Writing-original draft preparation: NK, SBD.

Writing-review and editing: NK, VB, JR, JB, JLC, SBD.

Supervision: JLC, SBD.

## Data Availability

The participant consent does not permit deposition of genetic data in a public repository. All the other data and materials supporting the findings of this study are available under a data transfer agreement from the corresponding authors on reasonable request.

## Ethics Approval and Consent to Participate

Written informed consent for participation was obtained from the patient’s family. Ethical approval for patient participation was granted by the Ethics Committee of Tehran University of Medical Sciences, Iran (protocol no. IR.TUMS.CHMC.REC.1397.004). The analysis of patient samples was approved by the Ministry of Higher Education and Research, France, under authorization DC 2024-6344. and The Rockefeller University Institutional Review Board, New York, United States, under protocol JCA-0699.

## Consent for Publication

Written informed consent for publication of the patient’s clinical data was provided by the family, with local IRB approval.

## Materials and methods

### Human participants

Both the patient and her living relatives studied here are alive and followed up in Iran. Written informed consent was obtained from all family members tested. Travel controls from Iran were provided. Experiments were performed in France, in accordance with local regulations. Whole-blood samples from the patient and her healthy relatives were collected into heparin-containing tubes.

### Sanger and whole-exome sequencing (WES)

Genomic DNA extracted from heparin-treated whole-blood samples from the patient and her healthy relatives was used for WES and verification by Sanger sequencing. WES was performed with the SureSelect Human All Exon V6 kit from Agilent on an Illumina sequencing platform. Homozygosity rates were estimated from genomic DNA, as previously described^30^. TNF exon 4 was amplified from genomic DNA with the DreamTaq DNA polymerase (#K1082; Thermo Fisher Scientific). Amplicons were then sequenced by the Sanger method with the Big Dye Terminator kit v3.1 (Thermo Fisher Scientific) and capillary electrophoresis (#A30469; Applied Biosystems 3500xL system, Thermo Fisher Scientific).

### Expression vectors

The constructs encoding TNF in a pCMV expression vector have been described elsewhere^5^. TNF c.673C>T was generated by PCR mutagenesis. The pGL4-NF-κB firefly luciferase and pRL-TK *Renilla* luciferase constructs have been described elsewhere^5^.

### Cell lines

HEK293T cells (human kidney epithelial cells; RRID:CVCL_0063) were maintained in Dulbecco’s modified Eagle’s medium (DMEM; Life Technologies). The culture medium was supplemented with 10% fetal calf serum (FCS; Sigma), 100 U/mL penicillin and 100 µg/mL streptomycin (GIBCO). HEK293T cells were cultured at 37°C under an atmosphere containing 10% CO_2_ in a humidified incubator. Cells were routinely screened for mycoplasma.

### Transient transfection

HEK293T were transfected with Xtreme Gene9 (Sigma), in accordance with the manufacturer’s instructions. The supernatant was harvested 24 h after transfection and used to stimulate cells in subsequent experiments and the cells were lysed for analysis.

### Cell stimulation

Where appropriate, cells were treated with the following inhibitors or ligands. Supernatants containing soluble TNF were generated by the transient transfection of HEK293T cells and were harvested 24 h post transfection. Recombinant human TNF was used at concentrations of 0.3, 3 and 30 ng/mL for the times indicated. For the detection of TNF, PMA and ionomycin were used at final concentrations of 40 ng/mL and 10^-2^ M, respectively, for 6 h. For the determination of intracellular TNF, GolgiPlug (Thermo Fisher Scientific; 1:1000) was added 1 h after the addition of PMA/ionomycin, and the sample was incubated for a further 5 h.

### Luciferase assay

HEK293T cells were transiently cotransfected, in technical triplicates, with the NFκB-firefly luciferase reporter construct and a constitutive *Renilla* luciferase construct in the presence of Xtreme Gene9, in accordance with the manufacturer’s instructions. We added supernatants containing soluble TNF to the cells 24 h post-transfection and incubated the cells for a further 24 h. Reporter activity was then measured with the Stop and Glo Reporter Assay System (Promega), according to the manufacturer’s instructions. Firefly luciferase activity was normalized against *Renilla* luciferase activity and is expressed as a fold-change relative to unstimulated cells (relative luminescence units; RLU). Luminescence was captured on a microplate reader (VICTOR Nivo™).

### RT-qPCR

Total RNA was isolated from cells with the Quick-RNA MicroPrep spin column kit (Zymogen, R1050), according to the manufacturer’s protocol. For cDNA synthesis, 10 ng of total RNA was used for random oligomer-primed reverse transcription with Superscript II Reverse Transcriptase (Invitrogen, 18064022). Quantitative RT-PCR was performed with the TaqMan Universal Master Mix (Applied Biosystems, 4305719) and Taqman probes for GUSB (4326320E) and TNF (Hs00174128_m1; Thermo Fisher Scientific).

### Immunoblotting

Cells were lysed directly in Laemmli sample buffer (Biorad) supplemented with NuPAGE™ sample reducing agent. The resulting cell lysates were centrifuged through a 20 µL filter tip to shred the DNA. The proteins present in the lysate were separated by sodium dodecyl sulfate-polyacrylamide gel electrophoresis (SDS-PAGE) in Tris-Glycine 12% polyacrylamide gels (Biorad) in Tris-glycine buffer (Biorad) under reducing conditions and the resulting bands were transferred electrophoretically to nitrocellulose membranes (Biorad) in a semi-dry transfer system. Membranes were blocked by incubation with 10% skim milk (w/v) in Tris-buffered saline (TBS) supplemented with 0.1% Tween 20 (TBS-T) for 30 min. Membranes were probed with primary antibodies (anti-TNF, ab1793, diluted in 3% v/v bovine serum albumin (BSA) in TBS-T at 1:1000) overnight at 4°C. Secondary anti-mouse antibodies (Biorad, #1706516) conjugated to horseradish peroxidase (HRP; diluted in 1% w/v BSA TBS-T at 1:5 000) were incubated with the membranes for 1 h at room temperature. The membranes were washed with TBS-T between antibody incubations and before visualization. Membranes were developed by enhanced chemiluminescence (West Atto; Sigma) with the ChemiDoc Touch Imaging System (Bio-Rad) and Image Lab software.

### Differentiation of monocyte-derived macrophages (MDMs)

CD14^+^ cells were isolated from peripheral blood mononuclear cells (PBMCs) with CD14 MicroBeads (Miltenyi Biotec, 130-050-201). GM-CSF-matured MDMs were generated with M1-Macrophage Generation Medium XF (PromoCell, C-28055).

### ROS production assay

Superoxide production by monocyte-derived macrophages (MDMs) was evaluated with the Superoxide Anion Assay Kit (Sigma-Aldrich, CS1000) according to the manufacturer’s instructions. In brief, 3 × 10^4^ MDMs were plated in a 96-well plate in the presence or absence of superoxide dismutase and stimulated with PMA (400 ng/mL). Luminescence was captured on a microplate reader (VICTOR Nivo™).

### Detection of intracellular TNF

PBMCs were isolated and dispensed into a 96-well round-bottomed plate at a density of 2 × 10^5^ cells per well. Cells were stimulated with PMA, ionomycin and GolgiPlug, as described above. After 6 h, cells were stained with fixable viability dye for 10 minutes at room temperature before fixation (eBioscience™ Foxp3 / Transcription Factor Staining Buffer Set) for 30 min on ice in the dark. The cells were then washed with permeabilization buffer (eBioscience™ Foxp3 / Transcription Factor Staining Buffer Set) and stained with anti-TNF-BV10 (Biolegend, 502950) and anti-IL1β-AF647 (Biolegend, 508208) overnight at 4°C in the dark. Cells were resuspended in FACS buffer (PBS supplemented with 2% FCS and 2 mM EDTA) and collected on a Gallios flow cytometer.

### Detection of soluble TNF

PBMCs were isolated and dispensed into a 96-well round-bottomed plate at a density of 2 × 10^5^ cells per well. Cells were stimulated with PMA and ionomycin as described above. The supernatants were collected after 6 h and evaluated with LEGENDplex Human Inflammation Panel 1.

### Mass cytometry on whole blood

Mass cytometry was performed on fresh heparin-treated whole-blood samples as previously described, with a customized panel^31^. All samples were processed within 36 h of blood sampling.

### Multiplex detection of anti-cytokine autoantibodies by flow cytometry

BD Cytometric Bead Arrays (BD CBA Flex system) were coated with a panel of recombinant human cytokines (10 µg; Biotechne) according to the manufacturer’s instructions (BD558556). Coupling was validated and the beads were then incubated for at least 2 h with serum or plasma samples from patients, positive controls and healthy donors. The beads were washed twice with phosphate-buffered saline (PBS; Gibco) supplemented with 0.005% Tween, and were then incubated with a PE-goat anti-human IgG antibody (SouthernBiotech C3923-S083E). After a final wash in PBS supplemented with 0.005% Tween, samples were acquired on an Agilent Novocyte NovoSampler Pro and the data were analyzed with FlowJo v.10.6.2 (Becton Dickinson).

### Serology (VirScan / IntelliFlex)

Plasma from 6 individuals (1 TNF variant carrier and 5 wild-type controls) were profiled on a 200-antigen IntelliFlex multiplex serology panel (Institut Pasteur, two plates). The instrument reports two reporter channels; the anti-IgG channel (RP1) was identified using the pooled human IgG (IgIV) control, which read 106-fold over blank in RP1 versus 1.4-fold in RP2, and the second channel was excluded from all analyses. Seropositivity was called as ln(MFI) above the antigen-specific, country-calibrated cutoff (France), a rule that reproduced 11,424 of 11,424 of the manufacturer’s own calls (100.00%). This defined a continuous, threshold-anchored score, d = ln(MFI) − cutoff, where d = 0 is the seropositivity threshold. Validated cutoffs existed for 56 of the 200 antigens (30 cosmopolitan, 26 travel-associated); the remaining 144, including all influenza, hepatitis, SARS-CoV-2 and vaccine antigens, were expressed only as robust z scores (median/MAD) against the 81 unrelated samples run on the same plates, and were never called positive or negative. Antibody breadth was defined as the number of seropositive antigens among the 30 cosmopolitan antigens. Groups were compared by two-sided Mann-Whitney tests, unadjusted. Because breadth is strongly age-dependent in this cohort (Spearman ρ = +0.58, P = 0.007, ages 4–86 years), comparisons were repeated on residuals of a linear fit of breadth on age. Per-antigen comparisons used Fisher’s exact test with Benjamini-Hochberg correction across the antigens tested. Analyses were performed in Python 3 (numpy, scipy, matplotlib).

### Quantification and statistical analysis

Results from technical triplicates are presented as the mean and standard deviation (S.D.). The statistical significance of differences between two groups was assessed in unpaired *t*-tests. All graphical data were prepared and analyzed in GraphPad Prism (Version 9). In all analyses, at the significance threshold was set at *p* = 0.05, with significance indicated as follows: p > 0.05 (n.s.), *p* < 0.05 (*), *p* < 0.01 (**), *p* < 0.001 (***), *p* < 0.0001 (****).

## Abbreviations

AR: Autosomal recessive
TB: Tuberculosis
*M. tb*: *Mycobacterium tuberculosis*
TNF: Tumor necrosis factor
GM-CSF: Granulocyte-macrophage colony-stimulating factor
MSMD: Mendelian susceptibility to mycobacterial disease
BCG: Bacillus Calmette–Guérin
IFN-γ: Interferon-gamma
ROS: Reactive oxygen species
LOF: Loss-of-function
CGD: Chronic granulomatous disease
CRP: C-reactive protein
PPD: Purified protein derivative
CT: Computerized tomography
CMV: Cytomegalovirus
EBNA: Epstein-Barr virus nuclear antigen 1
HAV: Hepatitis A virus
HSV: Herpes simplex virus
VCA: Viral capsid antigen
VZV: Varicella zoster virus
CADD: Combined annotation-dependent depletion
LOE: Loss of expression
PBMC: Peripheral blood mononuclear cells
PMA: Phorbol 12-myristate 13-acetate
MDM: Monocyte-derived macrophages
DHR: Dihydrorhodamine
TEMRA: T effector memory re-expressing CD45RA
MAIT: Mucosal-associated invariant T
NK: Natural killer
DC: Dendritic cell

